# Sustained patient use and improved outcomes with digital transformation of a COPD service: RECEIVER trial and DYNAMIC-SCOT COVID-19 scale-up response

**DOI:** 10.1101/2022.04.04.22273427

**Authors:** A Taylor, A Cushing, M Dow, J Anderson, G McDowell, S Lua, M Manthe, S Padmanabhan, S Burns, P McGinness, DJ Lowe, C Carlin

## Abstract

**Introduction:** LenusCOPD has been co-designed to enable digital transformation of COPD services for proactive preventative care. Patient-facing progressive web application, clinician dashboard and support website integrate patient-reported outcomes (PROs), self-management resources, structured clinical summary, wearable and home NIV data with asynchronous patient-clinician messaging. We commenced the implementation-effectiveness observational cohort RECEIVER trial in September 2019, with the primary endpoint of sustained patient usage and secondary endpoints including admissions, mortality, exacerbations, service workload and quality of life. We paused recruitment in March 2021 and provided LenusCOPD as routine care in the “DYNAMIC-SCOT” COVID-19 response service scale-up.

**Methods:** 83 RECEIVER trial participants and 142 DYNAMIC-SCOT participants had completed minimum 1 year follow-up when we censored data on 31^st^ August 2021. We established a control cohort with 5 patients matched per RECEIVER participant from de-identified contemporary routine clinical data.

**Results:** Sustained patient app utilisation was noted in both cohorts. Median time to admission or death was 43 days in control, 338 days in RECEIVER and 400 days in DYNAMIC-SCOT participants who had had a respiratory-related admission in the preceding year. The 12-month risk of admission or death was 74% in control patients, 53% in RECEIVER and 47% in the DYNAMIC-SCOT sub-cohort participants. There was a median of 2.5 COPD exacerbations per patient per year with stable quality of life across follow up and a manageable workload for clinical users.

**Conclusions:** A high proportion of people continued to use the co-designed LenusCOPD application during extended follow-up. Outcome data supports scale-up of this digital service transformation.

**Key messages:** 

**What is the key question?:** Can sustained patient interaction and improved patient outcomes be achieved with digital transformation of a COPD service?

**What is the bottom line?:** Participants continue to use the LenusCOPD patient app, with an average of 3-3.5 interactions per person per week sustained >1-year post-onboarding. COPD- related hospital admissions and occupied bed days were reduced following LenusCOPD onboarding in participants with a history of a severe exacerbation in the previous year, with a median time to readmission of 380 days compared with 50 days in a contemporary matched control patient cohort.

**Why read on?:** Feasibility and utility results support scale-up adoption of these digital tools, to support optimised co-management of COPD and other long-term conditions within a continuous implementation-evaluation framework. This will establish a test-bed infrastructure for additional innovations including artificial intelligence-insights for MDT decision support.

## Introduction

Chronic obstructive pulmonary disease (COPD) affects over 1.2 million people in the UK. COPD exacerbations account for one in eight UK hospital admissions, with a projected annual cost to NHS UK of £2.5bn by 2030. People with COPD prioritise the avoidance of exacerbations and resultant hospitalisations(1) The Patient Charter for COPD (March 2021), called for proactive, preventative management to reduce the risk of exacerbations and premature death(2). Strategies and services that support guideline-based care and self-management of COPD have been shown to reduce exacerbations and hospitalisations, improve health-related quality of life measures, improve long term outcomes and decrease healthcare and social costs(3–7). However, availability, accessibility, uptake and delivery of these interventions are highly variable(8).

Digital transformation with co-designed digital tools offers the opportunity to expand accessibility and engagement with these COPD interventions(9,10). Positive results have been seen from individual studies which have used internet/app based digital self-management interventions for COPD(11–13). Other studies have shown neutral or negative impact. The format and delivery of the interventions are variable, extensive heterogeneity between studies and high risk of bias means that systematic reviews have been unable to collate evidence of significant, persisting benefit of digital interventions(14).

Access and familiarity with digital interventions has been accelerated by the COVID- 19 pandemic, including in older adults(15). Improved usability and enhanced data-sharing enabled by cloud-based computing presents opportunities to improve on previous digital COPD interventions. Emerging innovations, including connected therapies (e.g., smart inhalers, home NIV), wearable sensors and AI-based analytics create possibilities for additional COPD care-quality improvements, if an infrastructure to implement and evaluate these can be established.

We conceived and commenced the ‘DYNAMIC’ (Digital innovation with remote data management and machined-learning algorithms to integrate care of high-risk COPD) project in September 2018. Patient participatory co-design with cycles of user experience testing was used to develop the “LenusCOPD’ patient and clinician web applications and support website. The anticipation was that service transformation based on this intervention would achive a reduction in respiratory-related admissions and occupied bed days in a high-risk COPD cohort, if sustained patient utilisation and supported co-management could be delivered. We commenced the ‘RECEIVER’ (remote management of COPD: evaluating the implementation of digital innovations to enable routine care) trial to evaluate this intervention in September 2019, with a primary endpoint of participant use of the LenusCOPD application(16). We selected an implementation-effectiveness observational cohort design, to allow for adaptations to the intervention and the implementation strategy based on planned interim evaluations(17).

We had to pause RECEIVER recruitment at month 6 of 12 at the start of the first UK COVID-19 lockdown (March 2020). Based on positive interim evaluations (sustained patient app usage) we elected to continue RECEIVER follow-up in recruited participants and adapt the trial resources to rapidly scale-up our provision of the LenusCOPD-based. The aim was to mitigate COVID-19 related service interruptions and enhance COPD co-management in vulnerable individuals. In May 2020, we sent an initial batch of text-message invitations to register with the service to people with COPD who were previously known to secondary care teams. This was supported by a social media awareness campaign run by NHS GG&C Communications Team. We continued with the evaluation framework established for RECEIVER in this ‘DYNAMIC-SCOT’ scale-up.

We can now report on the primary outcome (patient utilisation of the app) and key 1-year secondary outcomes for the people who were recruited to RECEIVER (Sept 2019 – March 2020) and in the first cohort of people who were onboarded to the service in May-August 2020 (DYNAMIC-SCOT scale-up implementation cycle 1) who had completed minimum 12 months of follow-up when data was censored on 31^st^ August 2021. To support these evaluations, we matched each RECEIVER participant with 5 contemporary control patients from NHS GG&C SafeHaven’s COPD dataset.

The objectives of this hybrid implementation-effectiveness trial and service evaluation were to determine the feasibility and utility of digital transformation of an NHS COPD service including

- establishing if participants would continue using the LenusCOPD patient application (feasibility, utilisation).
- determining the impact of the digital transformation on outcomes including COPD-related hospital admissions, participant mortality, community-managed COPD exacerbations, quality of life, and measurements of clinician service workload.

## Methods

### Study design, COVID-19 response, participants

RECEIVER is a prospective observational cohort hybrid implementation and effectiveness study, performed according to the UK Policy Framework for Health and Social Care Research. All recruited participants provided written informed consent. The protocol is published(16).

People with COPD attending secondary care in NHS GG&C were screened for eligibility. Inclusion criteria: >18 years of age; confirmed diagnosis of COPD (GOLD 2020)(7); a severe COPD exacerbation in the previous 12 months and/or chronic hypercapnic respiratory failure or sleep disordered breathing meeting established criteria for home NIV/CPAP treatment; personal or close contact with daily smartphone, tablet or desktop computer internet access to a web browser; able to give informed consent.

Inclusion criteria were retained for the DYNAMIC-SCOT scale-up, with the requirement for a severe COPD exacerbation in previous year and/or chronic respiratory failure inclusion criteria removed. Clinical team vetted patient-triggered applications to the service, to ensure COPD diagnosis and residence within NHS GG&C.

People were excluded from RECEIVER and DYNAMIC-SCOT if they had a communication barrier precluding the use of the COPD digital service.

### Intervention

The service consists of a patient app, a clinician app/dashboard and support website (Figure 1), with the same components utilised by both RECEIVER and DYNAMIC-SCOT scale-up participants. Additional information on “how it works” is available at https://support.nhscopd.scot. Further details of the intervention components, data processing and data storage are in the supplementary methods.

**Figure 1.**
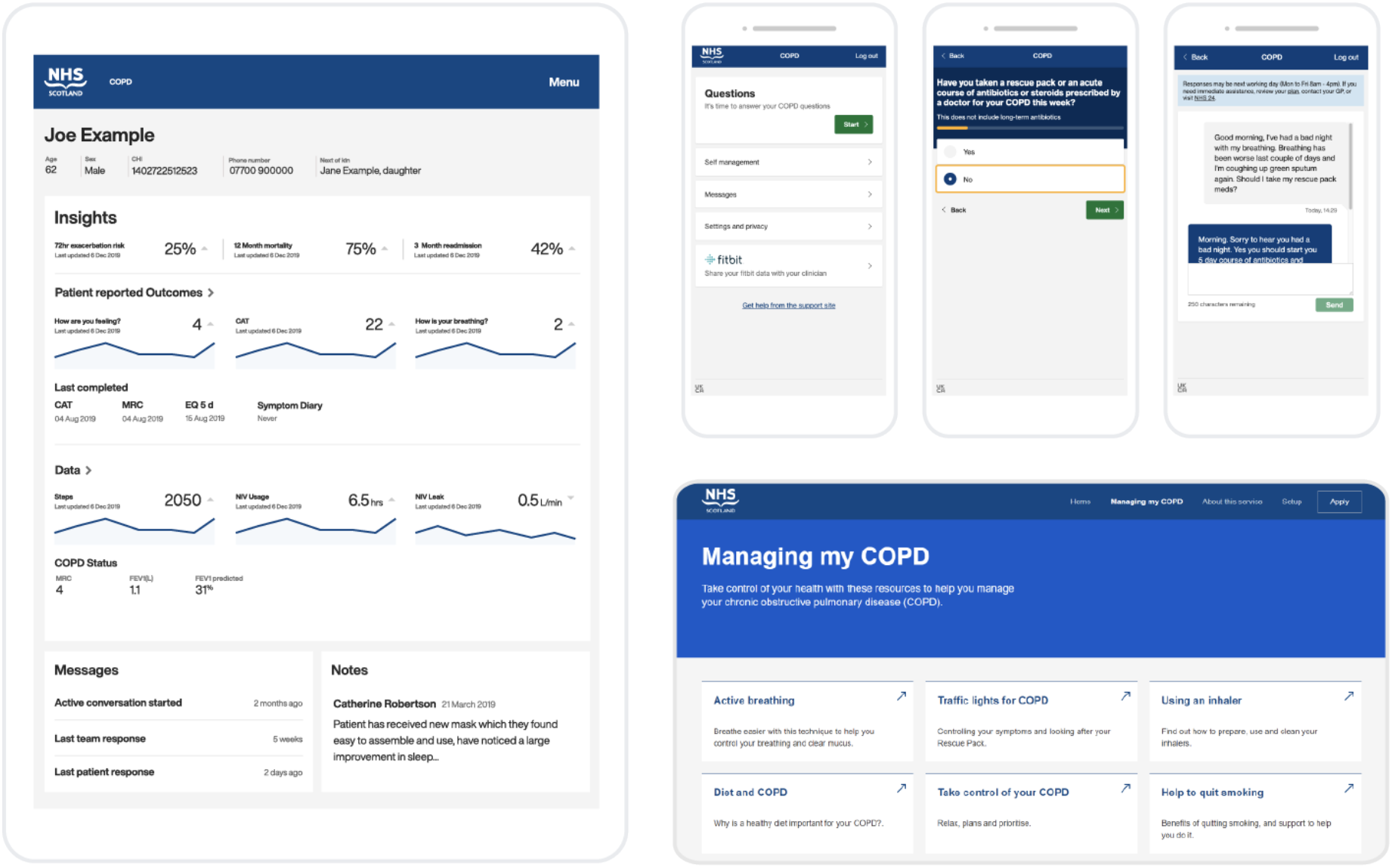
COPD digital service components. LenusCOPD clinician dashboard front page with sample data provides key data for “at a glance” review of patient status. Key patient app screens - opening page, weekly PRO question for COPD event detection, sample patient-clinician messaging with banner noting service purpose and emergency contacts – are included. Support website (https://support.nhscopd.scot) includes registration page for patient app, service explanation videos and COPD support content including self- management videos.

### Primary and Secondary outcome measures

The primary endpoint for RECEIVER trial was the proportion of enrolled participants with high-risk COPD who utilised remote management in a digital service model. Daily COPD assessment test and symptom diary questions, weekly MRC and healthcare episode questions, and monthly quality of life questions (EQ-5D-5L questionnaire) were captured in the LenusCOPD patient app. We pre-planned 3- monthly interim evaluations, with a target submission averaging >1 PRO set per participant per week as a benchmark, with the option to adapt the implementation strategy or intervention if required.

Supplementary table 1 summarises outcome measures planned in the RECEIVER trial and DYNAMIC-SCOT scale-up. Secondary outcomes reported here are respiratory-related hospitalisations, community-managed exacerbations reported in the patient app (detailed definitions in supplementary material), mortality, quality of life, clinician app user time and patient-clinician messaging volume. FitBit device wearable physiology, home NIV data via integration with the ‘AirView’ platform (ResMed) and qualitative data were collected in a proportion of RECEIVER trial participants. Analyses of these datasets, the detailed PRO insights and the associated AI model development are ongoing and will be reported subsequently.

**Table 1.** Baseline characteristics * n = 134, ** n = 45, ^ n = 133, ^^ n = 44

|  | RECEIVER | Scale-Up | Scale up<br>(admission<br>previous year) | Control |
| --- | --- | --- | --- | --- |
| Number of patients | 83 | 142 | 47 | 405 |
| Age at onboarding, mean (SD), years | 64.4 (9.31) | 65.9 (9.25) | 65.0 (10.0) | 64.3 (8.92) |
| Percentage female | 63.9 | 54.9 | 57.5 | 64 |
| n admissions previous year, mean (SD) | 2.46 (2.27) | 0.61 (1.21) | 1.85 (1.46) | 2.34 (2.56) |
| Smoking Status, % |  |  |  |  |
| Former Smoker | 70 | 78 | 79 |  |
| Current Smoker | 30 | 20 | 19 |  |
| FEV1% Predicted, mean (SD) | 47.89 (20.79) | 57.1 (24.75)* | 50.4 (22.2)** |  |
| FEV1/FVC Predicted, mean (SD) | 0.46 (0.14) | 0.49 (0.15)^ | 0.47 (0.15)^ |  |
| Baseline CAT mean (SD) | 23.22 (6.58) | 23.17 (8.06) | 25.28 (7.50) |  |
| Baseline MRC mean (SD) | 3.67 (1.19) | 3.41 (1.17) | 3.89 (1.18) |  |
| Highest Eosinophil Count, mean (SD) | 0.64 (1.10) | 0.50 (0.38) | 0.60 (0.45) |  |
| Triple combination inhalers (LABA+LAMA+ ICS), % | 80 | 62 | 72 |  |
| Previous pulmonary rehabilitation, % | 24 | 16 | 17 |  |
| NIV therapy, % | 29 | 3 | 6 |  |
| Home oxygen therapy, % | 37 | 16 | 32 |  |
| Comorbidities, % |  |  |  |  |
| Osteoporosis | 13 | 11 | 0 |  |
| Ischaemic Heart Disease | 8 | 25 | 21 |  |
| Obstructive Sleep Apnoea | 12 | 6 | 15 |  |
| Diabetes | 11 | 9 | 4 |  |
| Asthma | 10 | 13 | 0 |  |
| Atrial Fibrillation | 10 | 7 | 4 |  |
| DVT/PTE | 4 | 4 | 2 |  |
| Cerebrovascular Disease | 0 | 6 | 6 |  |
| Bronchiectasis | 2 | 8 | 6 |  |
| Pulmonary Hypertension | 1 | 0 | 0 |  |
| Pneumothorax | 2 | 4 | 6 |  |
| Pulmonary Fibrosis | 0 | 1 | 2 |  |
| Lung Cancer | 1 | 4 | 6 |  |

### Data collection

Baseline and follow-up demographic and physiological data from electronic health records is recorded in the LenusCOPD clinician dashboard. The index date for study participants is defined as the date on which they completed registration with LenusCOPD and had access to patient web application.

### Control cohort

NHS GG&C SafeHaven has a detailed de-identified dataset of demographic, laboratory, prescribing, hospital admission and mortality data. We followed relatable evaluations using routine clinical data by establishing a control cohort containing 5 patients matched to each RECEIVER trial participant(18). We selected matching criteria of diagnosis of COPD, age, sex and an index respiratory-related hospital admission within the 14-day window of the reference trial participant’s index date. We excluded duplicate control patients or those who had been subsequently onboarded to the COPD digital service.

### Sample size calculation

There is no reference data on uptake or sustained usage of a COPD patient app to allow calculation of a sample size. We set a screening target of 400 patients over 12-month period for the RECEIVER trial, based on previous experience(19).

### Statistical analysis

Survival to admission, death, and admission or death was estimated in the RECEIVER, DYNAMIC-SCOT scale-up, and control cohorts using Kaplan-Meier survival analysis with log rank test. To compare survival between cohorts, hazard ratios (HRs) and 95% lower/upper confidence intervals (CIs) were calculated using Cox regression. Wilcoxon signed-rank tests were used to compare the number of recorded admissions and occupied bed days in the 12-month period before and after index date. The significance and effect size of observed changes was determined for each cohort. Statistical analyses were performed using R Studio version 4.0.5 and GraphPad Prism version 9.3.1, with significance assessed at the 0.05 level.

### Patient and Public Involvement

Patients and members of the public were involved throughout the planning and conduct of this research. Semi-structured interviews were conducted by the DYNAMIC development team to understand patient’s experience of their condition and allow iterative co-design of the LenusCOPD tools. Pre-study testing yielded an average system usability score of 80/100, indicating high usability and learnability. The research questions and RECEIVER study design were informed by insights gained from these interactions and from published data on the priorities of people with COPD.

## Results

### Participants

The participant flow diagram details the recruitment and invitation strategies, number of people screened, recruited, commenced on intervention and who withdrew or died in the RECEIVER trial and in implementation cycle 1 of the DYNAMIC-SCOT service-scale up (Figure 2). Screening was ahead of target when we paused RECEIVER recruitment.

**Figure 2.**
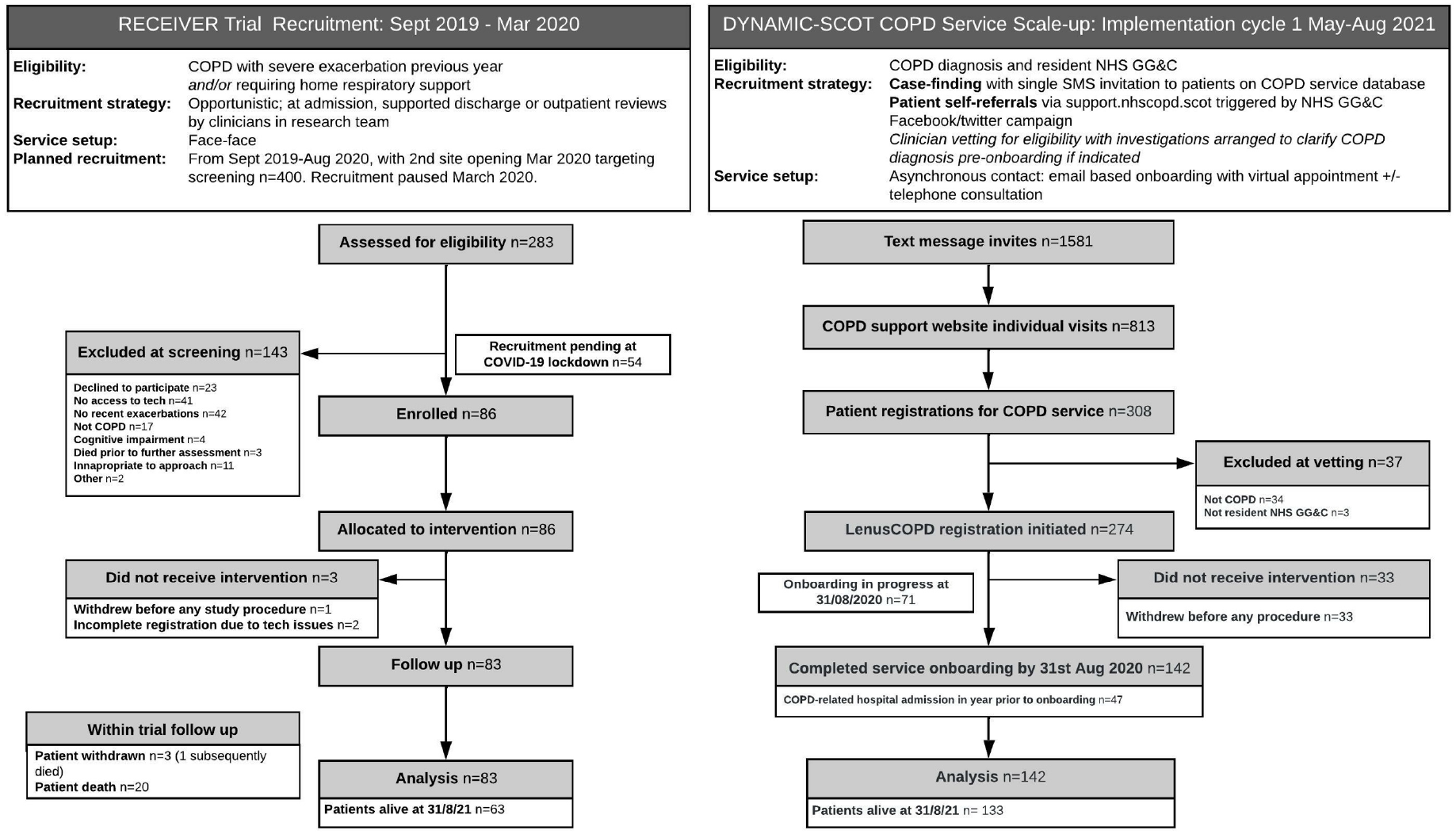
Participant flow diagram.

Baseline characteristics are summarised in table 1. People who commenced the intervention via the DYNAMIC-SCOT scale-up implementation had lower current smoking rates, less severe airflow obstruction, reduced number of hospitalisations in the previous year, reduced home oxygen or home NIV therapy requirement, lower rate of previous pulmonary rehabilitation completion, lower rates of osteoporosis and higher rates of bronchiectasis and ischaemic heart disease compared to RECEIVER participants. Baseline symptom burden (CAT and MRC scores) was similar for the cohorts. There was more severe airflow obstruction and higher rates of home oxygen therapy use in the sub-cohort of DYNAMIC-SCOT participants who had had a respiratory-related hospital admission in the year prior to onboarding. Based on available data, the control cohort appears well matched to the RECEIVER cohort.

73% of RECEIVER and 68% of DYNAMIC-SCOT participants were resident in the most deprived SIMD 1 and 2 quintiles, mirroring distribution of COPD burden in NHS GG&C and suggesting equality of access with this intervention (Supplementary Figure 1).

### Primary outcome: patient app utilisation

RECEIVER participants completed an average of 3.5 daily PRO sets per patient per week across the duration of follow up. DYNAMIC-SCOT participants completed an average of 3 daily PRO sets per patient per week across the duration of follow up (Figure 3A and 3B).

**Figure 3.**
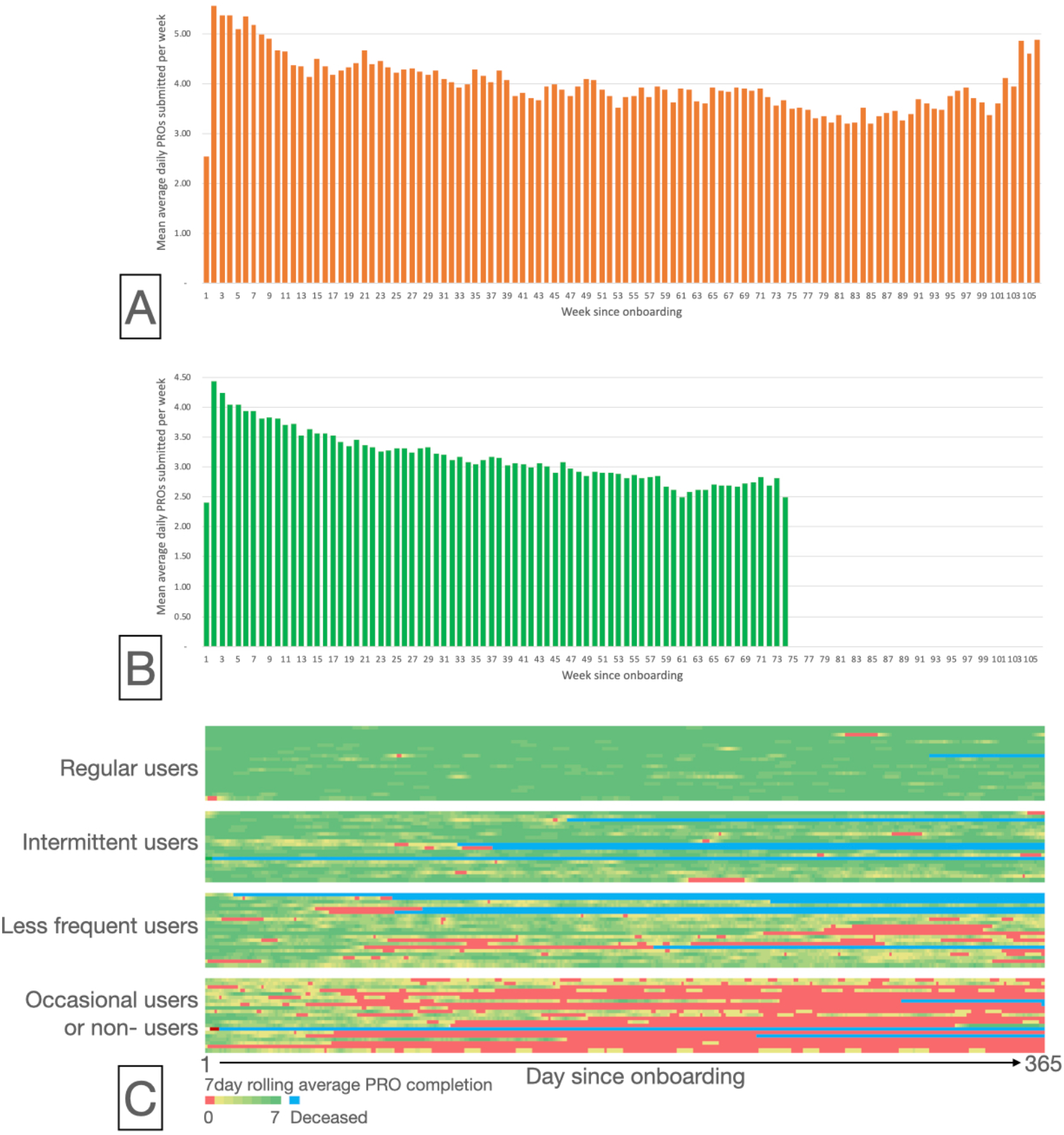
Patient reported outcome completion. A: Average of 3.5 daily patient reported outcomes are submitted per participant per week post onboarding in the RECEIVER cohort, with utilisation sustained during follow-up to week 106. Week 1 is a partial week for the majority of participants. B: Average of 3 daily PROs are submitted per participant per week post onboarding in the DYNAMIC-SCOT cohort, with utilisation sustained during follow-up to week 74. Week 1 is a partial week for the majority of participants. C: Per participant 7-day rolling average daily PRO completion heatmap at 1 year of follow up. Each row represents an individual RECEIVER trial participant. Participants are grouped by quartiles of the mean 7 day rolling average completion over 1 year.

80% of participants in RECEIVER and 60% of people in DYNAMIC-SCOT scale-up completed at least 1 PRO set per week on >50% of follow up weeks. There were no differences in completion rates between daily, weekly or 4-weekly PRO question sets (Supplementary Figure 2).

An exploratory evaluation of individual patterns of utilisation for participants in the RECEIVER trial was undertaken, with calculation of the average number of daily PRO sets completed in the previous week (7day rolling average). This is presented as a heatmap divided by quartiles of overall usage across 1 year of follow-up (Figure 3C). 39/83 participants paused their use of the intervention for >1 week, with the majority (24/39) resuming regular or intermittent interaction with the patient app across this follow-up period. There were no significant differences in baseline characteristics between these 4 groups (supplementary table 2).

**Table 2.** 12-month survival analyses: respiratory-related admissions and all-cause mortality.

|  | Control | RECEIVER | Scale-up<br>(admission in previous year) | Scale-up<br>(no admission in previous year) |
| --- | --- | --- | --- | --- |
| **Median time to event (days, interquartile range)** |  |  |  |  |
| Admission or death | 43 (4 - 284) | 338 (74 - 596) | 400 (161 - 450) | n/a |
| Death | n/a | n/a | n/a | n/a |
| Admission | 50 (2 - 236) | 380 (58 - 596) | 400 | n/a |
| **12-month respiratory-related admission or mortality** | 74.30% | 53% | 26.80% | 46.80% |
| **12-month mortality** | 32.60% | 16.90% | 8.50% | 5.3% |
| **12-month respiratory-related admission** | 66.20% | 47.00% | 26.10% | 46.8% |
| **Survival analyses: admission or death** | **Unadjusted hazard ratio (95% confidence intervals)** |  |  |  |
| Control vs RECEIVER | 0.47 (0.34 - 0.63) | p<0.0001 |  |  |
| Control vs Scale-up (previous admission) | 0.39 (0.26 - 0.59) | p<0.0001 |  |  |
| Control vs Scale-up (no admission) | 0.13 (0.07 - 0.2) | p<0.0001 |  |  |
| **Survival analyses: death** | **Unadjusted hazard ratio (95% confidence intervals)** |  |  |  |
| Control vs RECEIVER | 0.59 (0.37 - 0.94) | p=0.02 |  |  |
| Control vs Scale-up (previous admission) | 0.26 (0.11 - 0.64) | p=0.002 |  |  |
| Control vs Scale-up (no admission) | 0.16 (0.07 - 0.35) | p<0.0001 |  |  |
| **Survival analyses: admission** | **Unadjusted hazard ratio (95% confidence intervals)** |  |  |  |
| Control vs RECEIVER | 0.47 (0.34 - 0.63) | p<0.0001 |  |  |
| Control vs Scale-up (previous admission) | 0.41 (0.27 - 0.62) | p<0.0001 |  |  |
| Control vs Scale-up (no admission) | 0.13 (0.07 - 0.21) | p<0.0001 |  |  |

### Time to respiratory-related admission or death

Survival data is summarised in Table 2. The median time to respiratory-related admission or death in the RECEIVER cohort was 338 days (IQR 73.5 – 596 days) and 400 days (IQR 161-450 days) in the subgroup of DYNAMIC-SCOT participants who had had an admission in the year prior to onboarding, compared with 43 days (IQR 4-284 days) in the control cohort (Figure 4A). Hazard ratio for admission or death in RECEIVER participants was 0.47 (95% CI 0.34-0.63, p<0.0001), and 0.39 (95% CI 0.26-0.59, p<0.0001) in DYNAMIC-SCOT participants who had had an admission in the year prior to onboarding.

**Figure 4.**
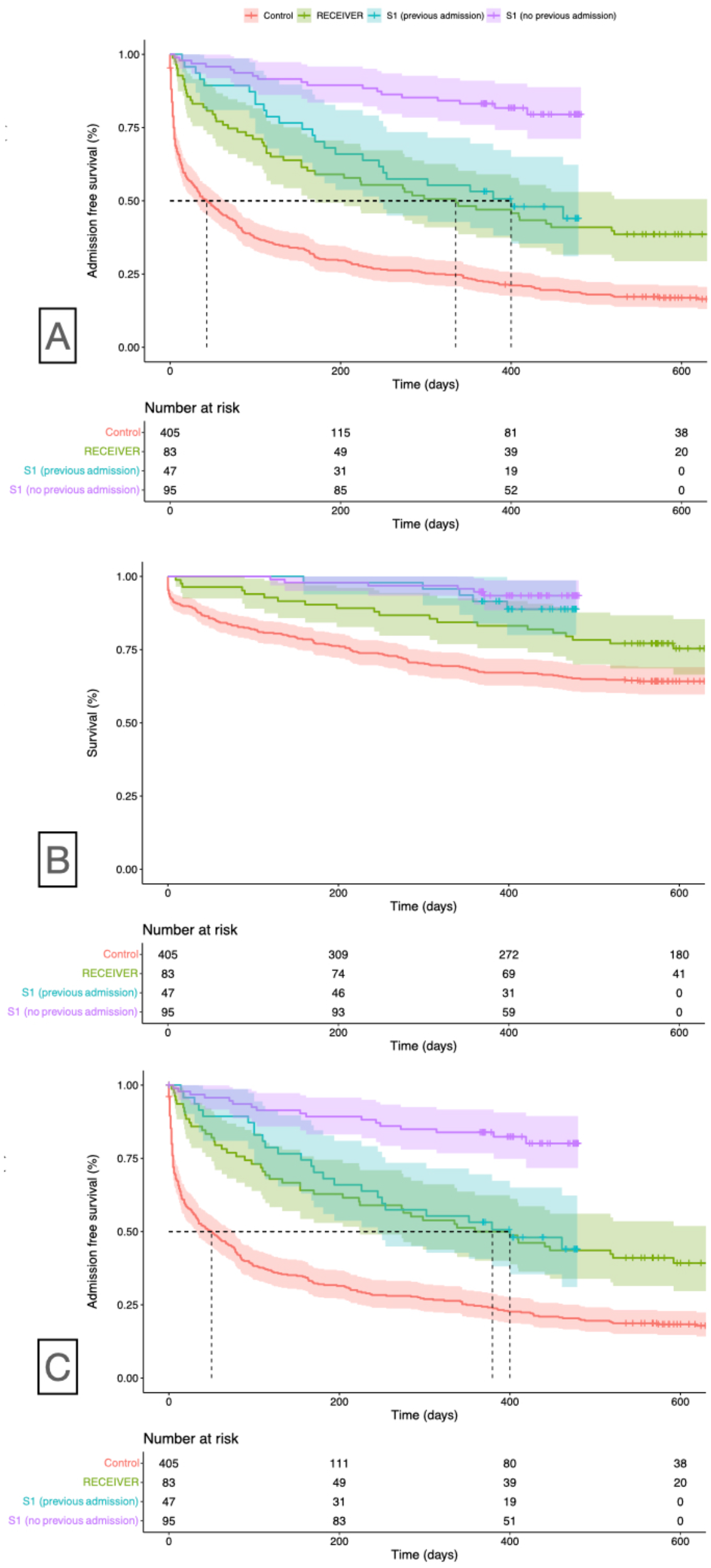
Kaplan-Meier survival plots of time to readmission or death (A), death (B) or readmission (C) from index/onboarding date until 31^st^ August 2021 in control, RECEIVER and DYNAMIC-SCOT cohorts (S1, subdivided by occurrence or absence of a respiratory-related admission in the year prior to onboarding).

Similar differences were noted with prolonged time to admission in the intervention cohorts when considering this endpoint alone (Table 2, Figure 4C).

No significance differences in time to admission survival metrics was seen in exploratory analyses conducted on RECEIVER participants stratified by 7-day rolling average PRO completion quartile or by SIMD category (Supplementary Figures 3 and 4).

### Mortality

12-month all-cause mortality was lower in the intervention cohorts than in the control cohort (Table 2; Figure 4B).

### Admissions and occupied bed days

Median respiratory-related admissions in the year prior vs the year-post index were reduced by 1 per patient per year in control participants.

Median respiratory-related admissions and occupied bed days in the year-prior vs the year-post index were reduced in RECEIVER and DYNAMIC-SCOT scale-up participants who had had an admission in the year prior to onboarding. Violin-box plot event rate distribution changes (Figure 5) and differences in effect sizes (Table 3) highlight the notable differences in these intervention cohorts versus controls.

**Table 3.** 12-month respiratory-related admission and occupied bed day event rate data in patients alive at 12-months post onboarding/index admission.

|  |  | Control | RECEIVER | Scale-up<br>(admission in previous year) | Scale-up |
| --- | --- | --- | --- | --- | --- |
|  |  | n=290 | n=69 | n=43 | n=133 |
| **Admissions** |  |  |  |  |  |
| Mean | Year Before | 2.06 | 2.2 | 1.84 | 0.59 |
|  | Year After | 1.51 | 1.01 | 1.21 | 0.5 |
| Median | Year Before | 2 | 2 | 1 | 0 |
|  | Year After | 1 | 0 | 0 | 0 |
| Mean Change |  | -0.55 | -1.19 | 0.63 | -0.09 |
| Median Change |  | -1 | -2 | -1 | 0 |
| Wilcoxon Signed-Rank Test (Significance) |  | p < 0.0001 | p < 0.0001 | p= 0.0030 | p= 0.192 |
| Wilcoxon Signed-Rank Test (Effect Size) |  | 0.327 | 0.594 | 0.503 | 0.127 |
| **Occupied bed days** |  |  |  |  |  |
| Mean | Year Before | 16.1 | 15.43 | 10.79 | 3.49 |
|  | Year After | 12.32 | 7.12 | 5.91 | 2.63 |
| Median | Year Before | 9 | 9 | 8 | 0 |
|  | Year After | 3 | 0 | 0 | 0 |
| Mean Change |  | -3.78 | -8.31 | -4.88 | -0.86 |
| Median Change |  | -6 | -9 | -8 | 0 |
| Wilcoxon Signed-Rank Test (Significance) |  | p < 0.0001 | p < 0.0001 | p= 0.0057 | p = 0.196 |
| Wilcoxon Signed-Rank Test (Effect Size) |  | 0.261 | 0.532 | 0.436 | 0.129 |

**Figure 5.**
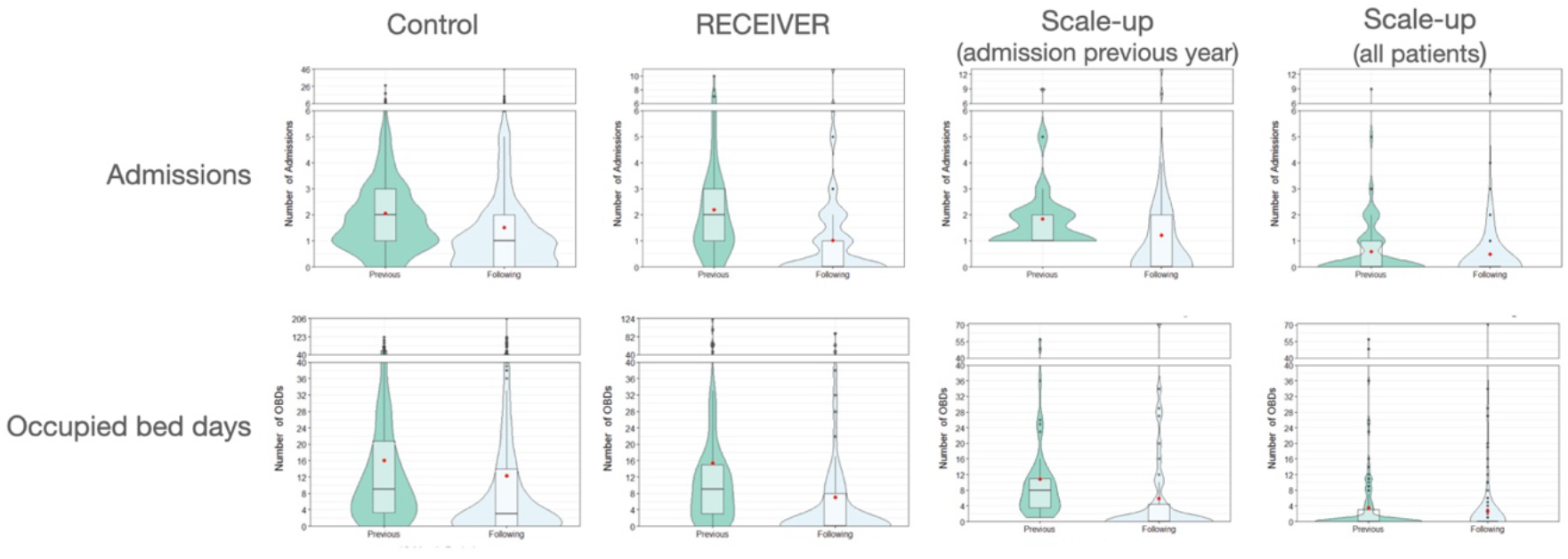
Violin boxplots summarising COPD-related hospital admission and occupied bed days in the year prior to and year following index/onboarding date in the 4 patient cohorts. Improved profiles are noted in the RECEIVER and Scale-up (admission previous year) intervention cohorts following onboarding compared to control cohort. Data are presented for patients alive at 12 months. No significant differences were noted in data distributions when including or excluding deceased patients (see supplementary content). Red dot = median. Horizontal bar = mean.

There were no differences in admission or OBD event rate distributions comparing the 4 subgroups of PRO completion or if analysis was restricted to patients alive at 12 months (supplementary tables 3 and 4).

### COPD exacerbations

RECEIVER participants reported a median of 2.5 and Scale-up participants a median of 2 community-managed exacerbations per patient per year in the 12- months post-service onboarding (Figure 6). A higher median of 4 community-managed exacerbations per patient per year was noted in the quartile of RECEIVER participants who interacted most frequently with the patient app (Supplementary Figure 5).

**Figure 6.**
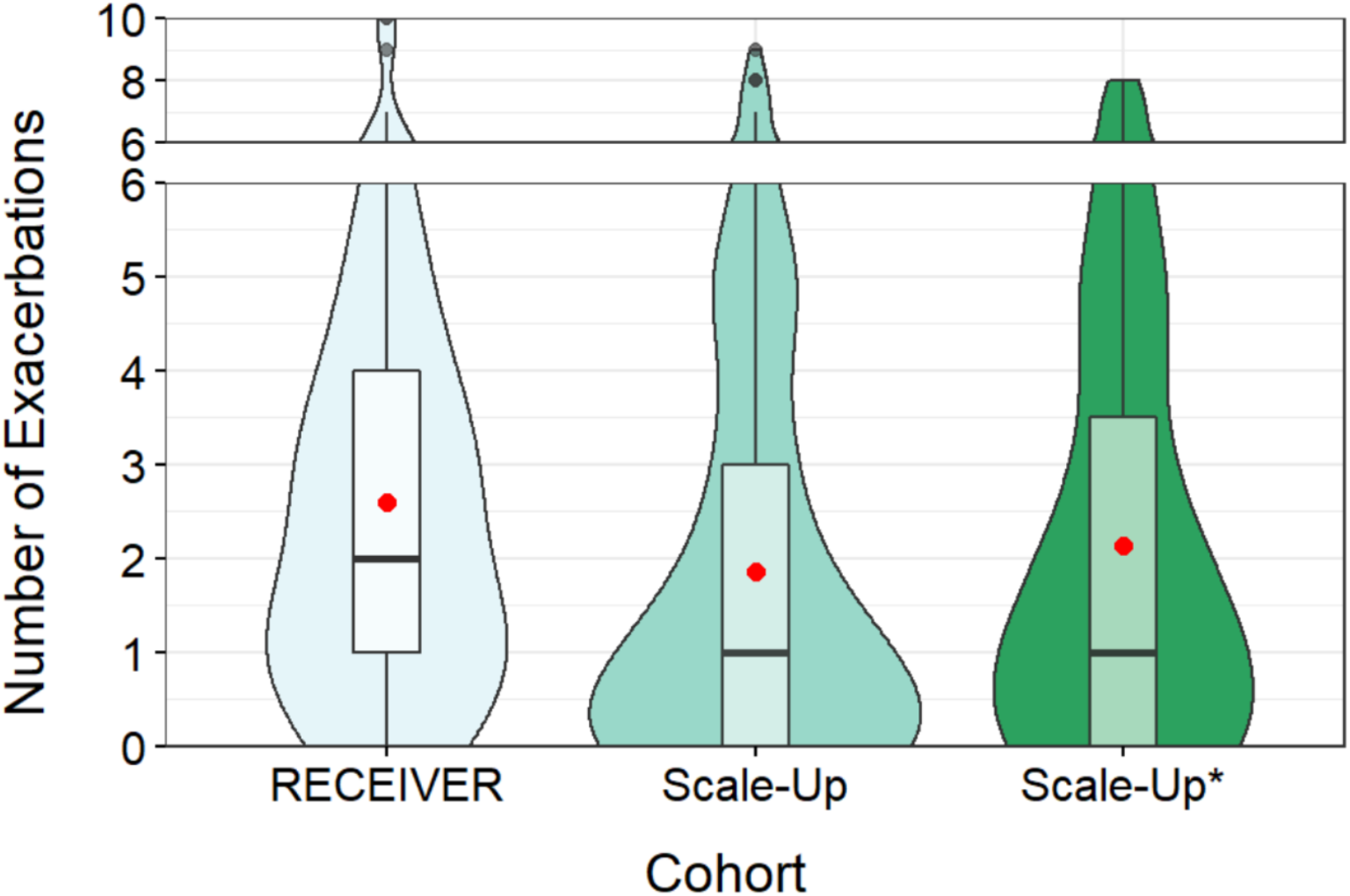
Violin boxplots summarising community-managed (clinician-assessed or self-treated) exacerbation events captured in the LenusCOPD patient app in the year following onboarding in the RECEIVER, DYNAMIC-SCOT scale-up intervention cohorts, and the sub-cohort of DYNAMIC-SCOT participants who had a respiratory-related admission in the year prior to onboarding (Scale-up*). Red dot = median. Horizontal bar = mean.

### Quality of life

We compared baseline and final CAT and EQ-VAS scores prior to censor date from participants who had completed >1 set of these PROs (Figure 7). There was a rise in median CAT score from 23/40 to 26/40 over the study follow-up period in both RECEIVER and scale-up cohorts (p= 0.008). There was no significant change in overall EQ-VAS scores across RECEIVER cohort, with a small reduction (median from 50 to 46.5) in Scale-up participants across follow-up. Variations in these and other captured PRO scores including per-patient level time-series with COPD events will be explored in subsequent analyses.

**Figure 7.**
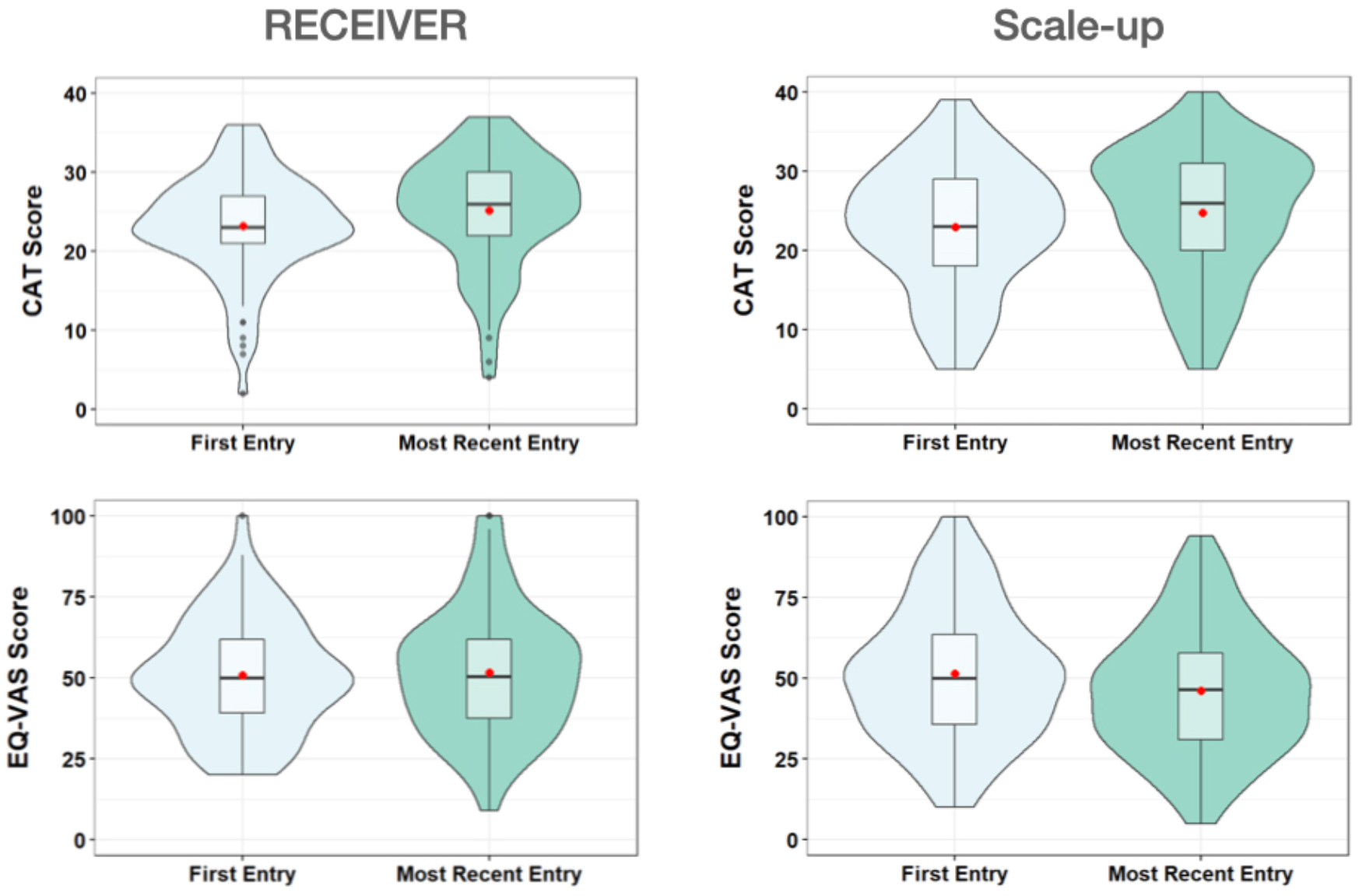
Violin boxplots summarising cohort distribution of CAT and EQ-VAS patient reported outcome scores from onboarding to most recent point completed in the RECEIVER and DYNAMIC-SCOT scale-up cohorts. Red dot = median. Horizontal bar = mean.

### Service workload

Clinical user login time averaged 4 hours per day Monday-Friday across the RECEIVER trial and DYNAMIC-SCOT scale-up period. 15-20 patient-clinician LenusCOPD messages are typically sent per working day within 4-5 individual participant conversations. Participant messaging frequency reduced steadily from 3-months post onboarding (Supplementary figure 5).

## Discussion

Sustained patient interaction with the co-designed LenusCOPD application was confirmed in the RECEIVER trial and the partnered DYNAMIC-SCOT scale-up service evaluation. Improved time to admission or death was noted in people onboarded to the LenusCOPD service compared with a contemporary control cohort. Reduction in post-onboarding annual respiratory-related hospital admission and occupied bed day rates was seen in RECEIVER participants and DYNAMIC- SCOT participants. These outcomes were obtained with an acceptable clinical team workload, an anticipated rate of community managed COPD exacerbations, and overall stable quality of life and disease impact metrics across the study follow up period.

These outcome findings are consistent with several previous studies of telehealth, remote-monitoring and supported self-management-based COPD service transformation, and contrast with individual studies and systematic reviews which have shown muted, no effect or a negative impact of comparable interventions on COPD events(11–14,20–23). Key differentiators for this LenusCOPD-enabled digital transformation include extensive pre-trial patient and clinician co-design (incorporating lessons learned from previous investigations), verified COPD diagnosis, daily prompts to complete PROs, asynchronous patient-clinician messaging, clinician option to activate individualised rescue pack advice in the patient app, and use of the service alongside routine care contacts rather than prespecified regular data reviews with triggered patient contact. The sustained participant utilisation of the patient app may also account for some of the apparent outcome benefits. Incorporating follow up beyond one year is also a strength compared to most other COPD intervention studies(20), and we will be able to report longer term follow-up subsequently.

Only 41 of 283 potential RECEIVER participants lacked technology access which was lower than anticipated and is improved compared with previous reports(24). Service uptake following a single unsolicited text message in the DYNAMIC-SCOT scale-up was also encouraging, as was the success of the remote-onboarding process. This demonstrates the appetite for supportive digital healthcare interventions such as LenusCOPD. There was a drop-off from website views to patient registrations, suggesting a need to augment trusted clinician contact and supporting resources. There was also a higher proportion of regular users and overall participant interactions in the RECEIVER cohort compared to the DYNAMIC- SCOT cohort, potentially reflecting different patient motivations. Exploratory survival and event-rate analyses in the RECIEVER cohort reassuringly showed no significant outcome differences with less frequent daily PRO completion, with a high proportion of participants with interrupted usage resuming interactions over the follow-up year. Whilst patients receive a text and email prompt to answer daily COPD questions, we didn’t prespecify any usage pattern for patient or clinical users. It is not possible to determine the relative benefit of regular participant interaction from specific component(s) of this multifaceted intervention. The ongoing RECEIVER analyses (supplementary table 1) will provide further insights on the patient-perceived benefits, motivations and barriers for usage and potential avenues for improvement.

The NOVELTY study has established that PROs and event rates provide better risk stratification than clinician-determined severity classification(25). The LenusCOPD- based intervention offers the facility for assured continuous capture of this patient data, with service and technical interoperability for triggered direct care and research evaluations. Implementing and evaluating derived AI-based actionable insights within a COPD MDT are a logical next step.

The service workload metrics reportable here are also encouraging and compatible with anticipated benefits of digital transformation and implementation of assistive technologies. Based on the positive RECEIVER/DYNAMIC-SCOT evaluations we will continue scale-up and adapt LenusCOPD to support COPD diagnostic evaluation, blended pulmonary rehabilitation service delivery, and scheduled review in primary care. An adoption playbook has been developed to support other clinical teams, and pilot adoption experience in an adjacent health board has been positive (C Yerramasu, NHS Lothian, personal communication).

The strengths of this study include the selection of an observational implementation-effectiveness methodology, which allowed us to pivot the implementation to assist COVID-19 pandemic response. The use of contemporary routine clinical data to derive a control cohort rather than a randomised design allowed us to maximise participant numbers using the intervention for the interim and final primary endpoint analyses. Whilst the lack of a randomised control arm is an important caveat, the control cohort derivation with an index date and outcome follow-up time period matched to RECEIVER participants reduces potential bias from seasonality and COVID-19 pandemic impacts on COPD event rates. Conclusions from this study have to be tempered by residual biases and confounders, including incomplete clinical information available for the control cohort and reduced integrity of matching for the control:DYNAMIC-SCOT comparison. Reduced event rates were seen in the control cohort across COVID-19 pandemic, this is in line with other investigations and suggests that this cohort is representative(26,27). The increased time to first events in the survival analyses and reduced median admission rates in the intervention cohorts provide a reassuring safety signal and encouraging hypothesis-generating utility data. The admission and community exacerbation event rates in the intervention cohorts provide further safety data: they are reduced when compared with historical NHS GG&C data, and similar to recent published data. It is therefore unlikely that reduced admission events in the intervention cohorts relate to delays from inappropriate community management, or that the intervention leads to unsupervised overuse of corticosteroid/antibiotic courses.

The reduced median occupied bed day rate profiles in the intervention cohorts are particularly encouraging. COPD is estimated to consume €48.4 billion annually in EU healthcare costs, with the majority of those costs arising from exacerbation and admission management(28). Scaling a service intervention which reduces respiratory-related occupied bed day rates in people with COPD should be highly cost-effective.

The Lenus-COPD based service transformation was co-designed with an experienced COPD multidisciplinary team and adopted “on top” of the team’s proven capabilities(29,30). COPD prevalence and burden is higher than population average in our predominantly urban health board, reflecting public health factors. The impaired survival and high admission event rates in our control cohort highlight a substantial persisting care-quality gap. We hope to address this this by scaling our digital transformation work in our own organisation, with continued evaluation and reporting. Our COPD digital service transformation strategy and evaluation framework should be adapted by different healthcare organisations and different territories, to reflect local services and clinical priorities.

## Conclusion

The data from the RECEIVER trial and DYNAMIC-SCOT COVID-19 response service scale-up support expansion of digital transformation of COPD services using co-designed tools such as LenusCOPD. The innovative implementation-effectiveness design with supporting contemporary matched routine clinical data allowed rigorous evaluation of an intervention to be combined with adjustments to the intervention or implementation strategy based on adoption experience, or if the adoption scenario changes. The LenusCOPD-based service transformation co-designed in the DYNAMIC project provides wide-ranging opportunities to improve our understanding of COPD and achieve the outcomes – reduced exacerbations and associated admissions – which people living with COPD most value.

## Data Availability

All data produced in the present study are available upon reasonable request to the authors

https://support.nhscopd.scot

## Ethics approval

Ethical approval for the RECEIVER trial was obtained from the West of Scotland Research Ethics Service, reference 19/WS/0072. Approval for use of de-identifed electronic healthcare record data stored within SafeHaven repository was granted by NHS GG&C Local Privacy and Advisory Committee, reference GSH/19/RM003. NHS GG&C Caldicott Guardian approval was obtained for the DYNAMIC-SCOT scale-up service evaluation.

