## Supplementary materials for "Sustained patient use and improved outcomes with digital transformation of a COPD service: RECEIVER trial and DYNAMIC-SCOT COVID-19 scale-up response"

|  | **Secondary outcome reported in this analysis** | **Secondary outcome measured but not analysed** |
| --- | --- | --- |
| Clinical outcomes | Mortality  Respiratory–related ED attendances Respiratory-related hospital admissions Hospital occupied bed days  Community-managed COPD exacerbations | Primary care visits Treatment uptake (smoking cessation, pulmonary rehab, vaccination, home oxygen, home NIV) |
| Impact of demographics |  | Gender Smoking status SMID Participation in pulmonary rehab in preceding 2 years Lung function measurements Highest eosinophil counts Inhaled therapies Additional COPD therapies Patient activation measures Modelled home air pollution exposure  Age |
| Platform usage | Average PRO completion rate Messaging utilisation (number of message threads) | Utilisation of self-management information (page views) |
| Patient-centred outcomes | Health-related quality of life (EQ-5D-5L), COPD assessment tool (CAT), MRC breathlessness score and symptom diary trends *(partial information reported here)* | Qualitative participant research (semi-structured user experience interviews) |
| Physiological measurements |  | Step count/Heart rate/Sleep (Fitbit device)  NIV- usage, therapy parameters |
| Healthcare cost analyses |  | Projected direct and indirect cost savings of high-risk COPD with digitally enabled remote-management  Development and implementation costs  Recurring costs for service model |

**Supplementary Table 1** RECEIVER trial secondary objectives, subdivided by those included in this report, and those for which analyses are ongoing.

### Supplementary methods

#### Intervention

The patient app is a progressive, responsive web-application, accessible through smart phone, tablet or computer. Participants are sent a daily text message and email reminder to complete standard patient-reported outcome (PRO) questionnaires to aid structured reflection on COPD symptoms. The app provides access to standardised self-management advice (‘traffic lights’), individualised rescue-pack advice (when activated by clinician) and patient-clinician asynchronous messaging for non-urgent advice regarding COPD self-management.

Registration to the service prompts clinician review and completion of structured clinical summary data in the web-browser based clinical dashboard. The structured content supports comprehensive review and optimisation of COPD management. The NHS GG&C COPD clinical team additionally use messaging facility and the rich dataset aggregated in the dashboard to support scheduled care and respond to patient support messages. The patient app has a banner noting that clinical team review and respond to messages Mon-Fri 8am-4pm, with signposting to alternative channels for emergency care.

Clinical events are captured in the clinician app via PROs (weekly question set establishes if an exacerbation has occurred, Figure 1), with intermittent systematic electronic health record (EHR) review by clinicians to input hospital admissions.

All data was stored within NHS GG&C Azure cloud tenancy with standard NHS data control, processing and security governance arrangements. Participant permission for data sharing is additionally captured within the patient app. Identity was linked by unique patient identifier. Analysis of data generated was undertaken by named researchers using a de-identified data feed from LenusCOPD database. Analysis of historical control data was undertaken by named researchers working in NHS GG&C SafeHaven trusted research environment.

#### Definition of respiratory-related hospital admission

Respiratory-related emergency department (ED) attendances and hospital admissions were collated by research team from review of electronic health records for RECEIVER and DYNAMIC-SCOT scale up cohort in the 12 months preceding onboarding to LenusCOPD service and admissions post onboarding to 31^st^ August 2021. Occupied bed days were counted in whole days from point of attendance to discharge; ED attendances of <24 hours were therefore counted as one day. Elective admissions were included if they related to optimisation of COPD management. Where there were uncertainties relating to the nature of an admission, consensus was achieved by discussion within the investigator team.

Respiratory-related ICD-10 diagnosis codes were applied to the ED attendance/admission data for the control cohort to acquire admission event and occupied bed day counts.

#### Definition of a community-managed COPD exacerbation based on responses to weekly PRO questions in LenusCOPD

Exacerbations were identified from a once weekly PRO question in the patient application which asked the user “have you taken antibiotics/steroids in the last week?” A ‘YES’ response was designed to be surrogate marker of an exacerbation in the previous week. To account for sustained symptoms and extended courses of therapy deployed by some patient’s criteria for refining which ‘YES’ responses to the weekly PRO were classed as new exacerbation events were developed and referred to as the PRO LOGIC criteria. The PRO LOGIC criteria for defining a new exacerbation event from a ‘YES’ response from the weekly PRO question were as follows:

• More than 5 weeks since previous ‘YES’ response = always considered a new exacerbation event.

• More than 2 weeks but less than 5 weeks since last ‘YES’ response: ONLY = new exacerbation event IF two consecutive ‘NO’ responses between the previous ‘YES’ response and ‘YES’ response in question.

LenusCOPD messaging, therapy and emergency clinical summary prescribing data was reviewed to allow clinician verification of a selection of community-managed exacerbations. Good correlation between ‘PRO LOGIC’ and clinician-verifiable events was noted.

### Supplementary results

| **Characteristic** | **Significance** |
| --- | --- |
| Age | 0.3318 (Kruskal-Wallis) |
| Sex | 0.4992 (Chi-Sqaure) |
| Smoking Status | 0.7093 (Chi -square) |
| No. of previous admissions | 0.9919 (Kruskal-Wallis) |
| SIMD quintile | 0.7684 (Chi-Sqaure) |
| FEV1% Predicted | 0.8428 (Kruskal-Wallis) |
| FEV1 - FVC Predicted | 0.9837 (Kruskal-Wallis) |
| Completed PR | 0.4029 (Chi square) |
| NIV User at Baseline | 0.3221 (Chi-square) |
| Home oxygen User | 0.7331 (Chi-Square |
| Baseline CAT | 0.4737 (Kruskal-Wallis) |
| Baseline MRC | 0.2728 (Kruskal-Wallis) |
| Highest Eosinophil Count | 0.6877 (Kruskal-Wallis) |
| Osteoporosis Prevalence | 0.1126 (Chi-Sqaure) |
| Ischaemic Heart Disease Prevalence | 0.6514 (Chi-Square) |
| Obstructive Sleep Apnoea Prevalence | 0.2690 (Chi-Square) |
| Diabetes Mellitus Prevalence | 0.1578 (Chi-Sqaure) |
| Asthma Overlap Prevalence | 0.7777 (Chi-Square) |
| Bronchiectasis Prevalence | 0.5515 (Chi-Sqaure) |
| Atrial Fibrillation Prevalence | 0.5542 (Chi-Square) |
| Heart Failure Prevalence | 0.7213 (Chi-Square) |
| Cerebrovascular Disease Prevalence | NA |
| Pulmonary Fibrosis Prevalence | NA |
| Pulmonary Hypertension Prevalence | 0.3861 (Chi-Square) |
| Previous Pneumothorax Prevalence | 0.5723 (Chi-Square) |
| Deep Vein Thrombosis or Pulmonary Thromboembolism Prevalence | 0.285 (Chi-Square) |
| Lung Cancer Prevalence | 0.3934 (Chi-Square) |

**Supplementary table 2** RECEIVER cohort baseline characteristics: differences by PRO-type. Chi-square tests and Kruskall-Wallis tests were used as appropriate to determine if there was a significant difference in the baseline characteristics of study participants in each of the PRO completion quartiles. P-values are shown, no significant differences were noted.

| **Events** | **Significance** |
| --- | --- |
| Number of Admissions (All) | 0.5286 (Kruskal-Wallis) |
| Number of Occupied Bed Days (All) | 0.4335 (Kruskal-Wallis) |
| Time to First Readmission (All) | 0.1624 (Kruskal-Wallis) |
| Number of Admissions (Year) | 0.6713 (Kruskal-Wallis) |
| Number of Occupied Bed Days (Year) | 0.3397 (Kruskal-Wallis) |
| Time to First Readmission (Year) | 0.4444 (Kruskal-Wallis) |

**Supplementary table 3** RECEIVER cohort clinical events: differences by PRO-type. Kruskal-Wallis tests were used to determine if there was a significant difference in the number of admissions and occupied bed days and time to first readmission between study participants in each of the four PRO completion quartiles. p-values are shown, no significant differences were noted.

All= All RECEIVER participants

Year = RECEIVER participants alive 365 days following service enrolment.

**Admissions**

|  | | **Control (n= 405)** | **RECEIVER (n= 83)** | **Scale-Up (n = 142)** | **Scale-Up (Previous Admission) (n = 48)** |
| --- | --- | --- | --- | --- | --- |
| **Mean** | **Year Before** | 2.34 | 2.46 | 0.61 | 1.88 |
|  | **Year After** | 1.53 | 1.19 | 0.61 | 1.33 |
| **Median** | **Year Before** | 2 | 2 | 0 | 1 |
|  | **Year After** | 1 | 0 | 0 | 0 |
| **Mean Change** | | -0.81 | -1.27 | 0 | -0.55 |
| **Median Change** | | -1 | -2 | 0 | -1 |
| **Wilcoxon Signed-Rank Test (sig.)** | | p < 0.0001 | p < 0.0001 | p = 0.883 | p = 0.0091 |
| **Wilcoxon Signed-Rank Test (Effect Size)** | | 0.389 | 0.572 | 0.0389 | 0.429 |

**Occupied bed days**

|  | | **Control (n = 405)** | **RECEIVER (n= 83)** | **Scale-Up (n= 142)** | **Scale-Up (Previous Admission) (n = 48)** |
| --- | --- | --- | --- | --- | --- |
| **Mean** | **Year Before** | 20.71 | 19.39 | 3.62 | 11.67 |
|  | **Year After** | 14.02 | 9.99 | 3.78 | 7.77 |
| **Median** | **Year Before** | 12 | 11 | 0 | 8 |
|  | **Year After** | 4 | 0 | 0 | 0 |
| **Mean Change** | | -6.69 | -9.4 | + 0.16 | -3.9 |
| **Median Change** | | -8 | -11 | 0 | -8 |
| **Wilcoxon Signed-Rank Test (sig.)** | | p < 0.0001 | p < 0.0001 | p = 0.711 | p = 0.0148 |
| **Wilcoxon Signed-Rank Test (Effect Size)** | | 0.287 | 0.489 | 0.0636 | 0.367 |

**Supplementary table 4** 12-month respiratory-related admission and occupied bed day event rate data (all patients)

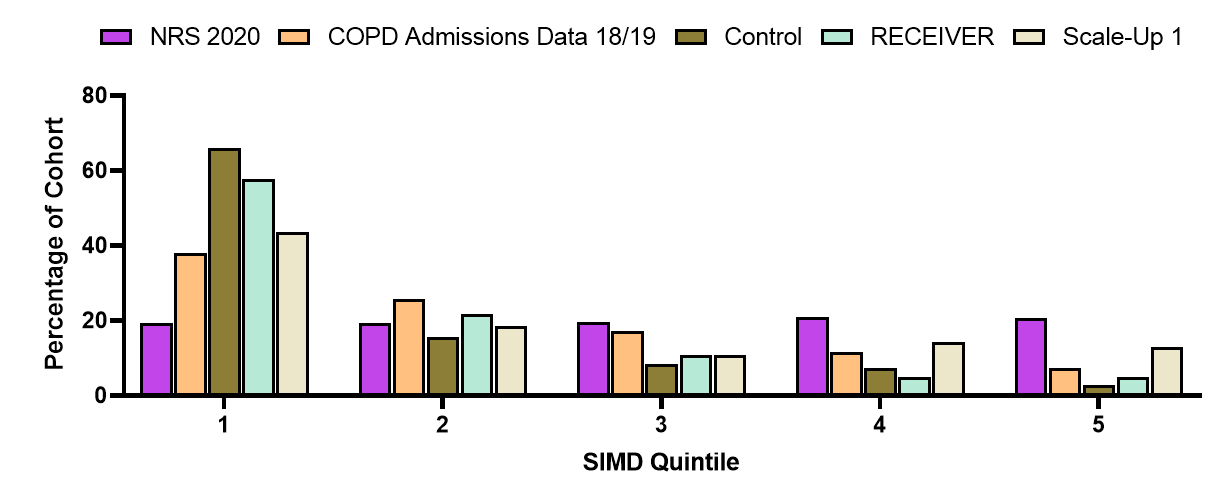

**Supplementary Figure 1** Distribution of residence by SIMD quintile 1 (most deprived) to 5 (least deprived).

NRS 2020 = SIMD distribution in Scottish population data.
COPD admissions data = from published Public Health Scotland Scottish Atlas of Variation 2018/19 <https://www.isdscotland.org/products-and-services/scottish-atlas-of-variation/view-the-atlas/respiratory.asp>

SIMD distribution of control cohort patients matches published data on COPD event rates in Scotland and the variation in care based on public health factors present in NHS GG&C.

>70% of patients onboarded to COPD digital service are resident in SIMD 1 and 2 postcodes, which is a higher proportion than Scottish distribution based on COPD related admissions. This reassures us that LenusCOPD service uptake is matching disease prevalence and equality access requirements in NHS GG&C.
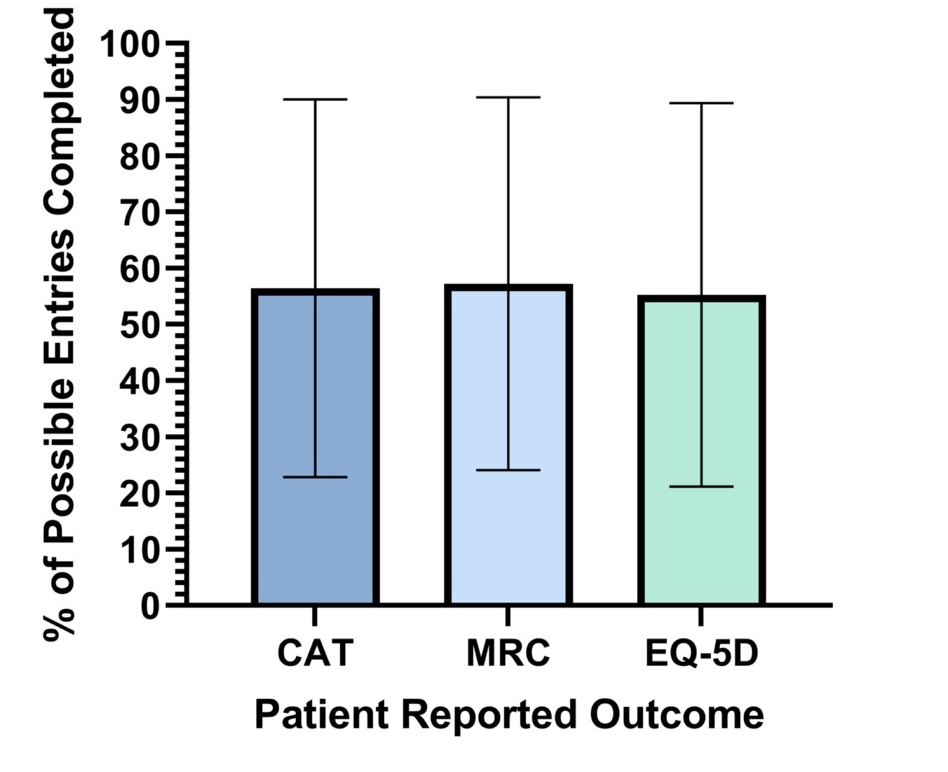

**Supplementary Figure 2** Mean percentage PRO completion by opportunity amongst the RECEIVER participants for daily CAT, weekly MRC and four-weekly EQ-5D-5L entries. Minimal variation is seen between the average completion rate, confirming that additional weekly and monthly questions did not appear to alter overall completion rates.

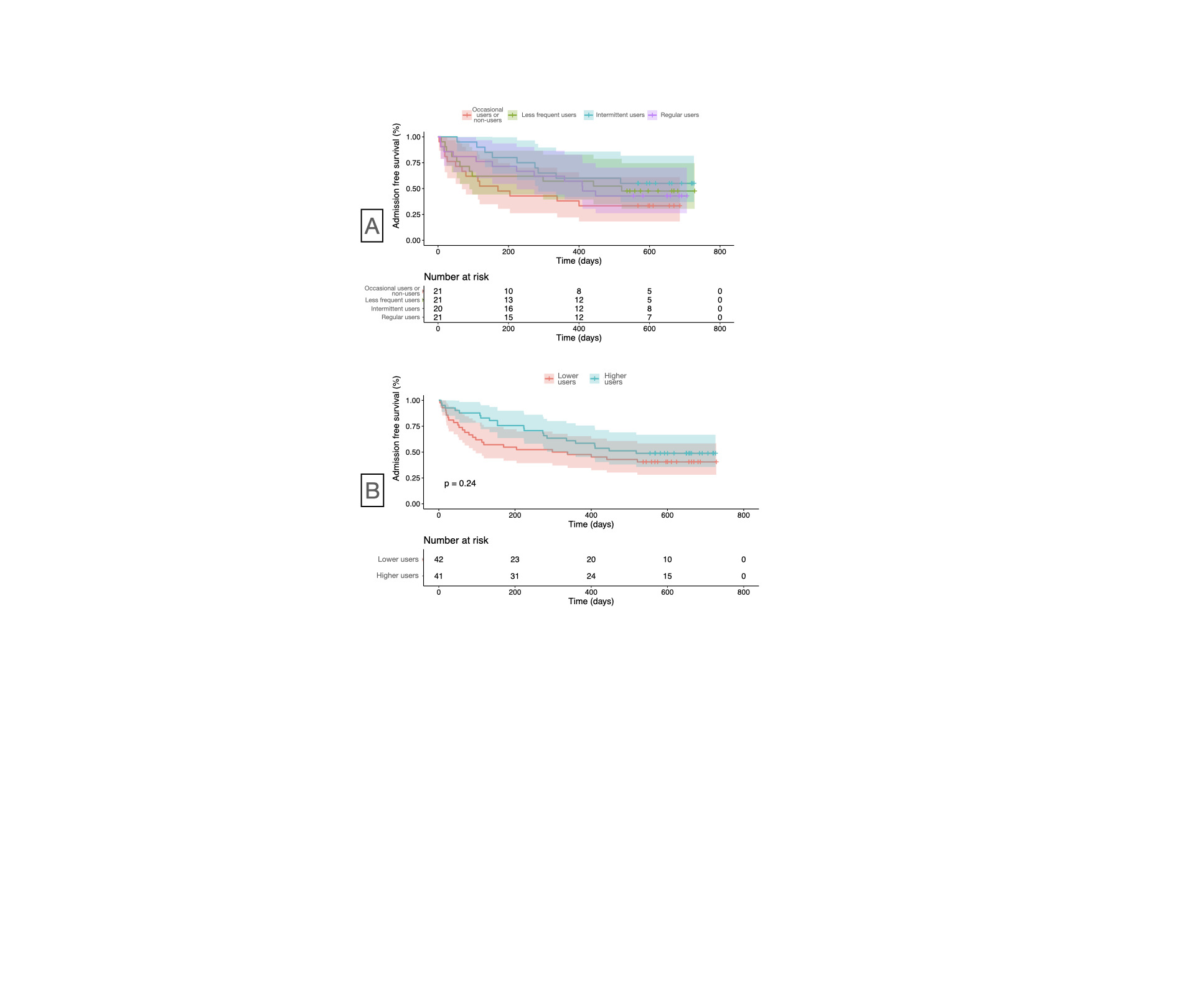

**Supplementary Figure 3** Kaplan-Meier survival plot showing no significant difference in time to admission from onboarding date until 31^st^ August 2021 in RECEIVER patients subdivided by PRO-completion quartiles (A). Non-significant trend to reduced time to admission is seen in RECEIVER participants subdivided by 2 highest usage and 2 lowest usage PRO-completion quartiles combined (B). See Figure 3C for PRO-completion data.

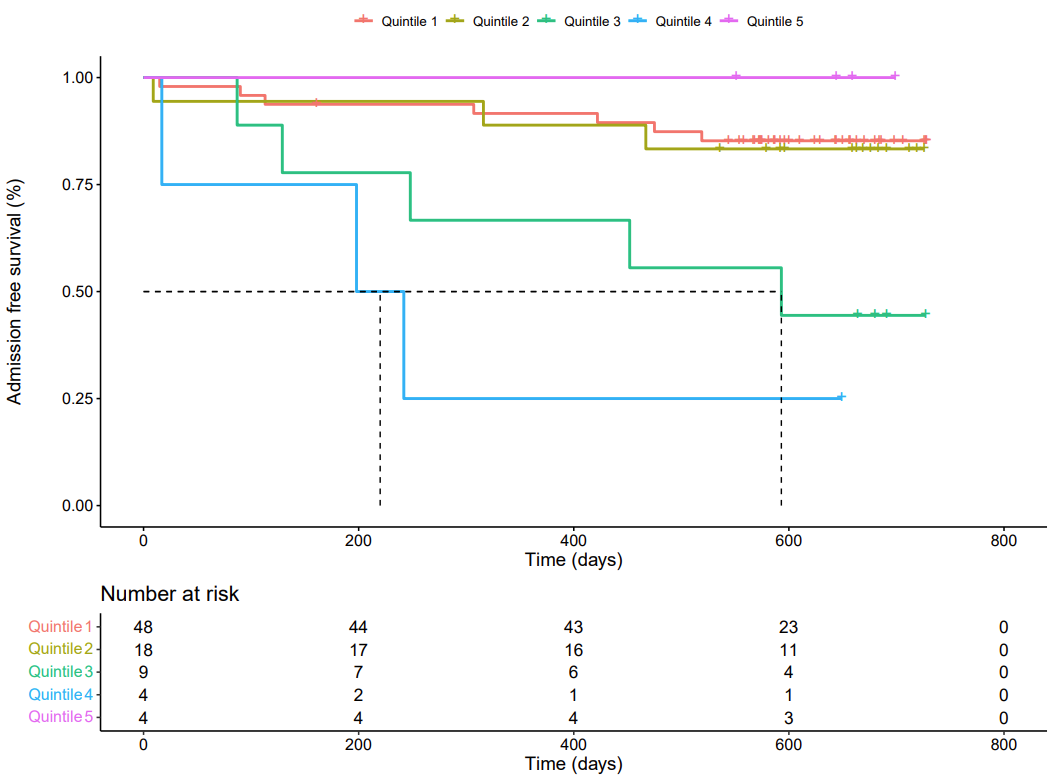

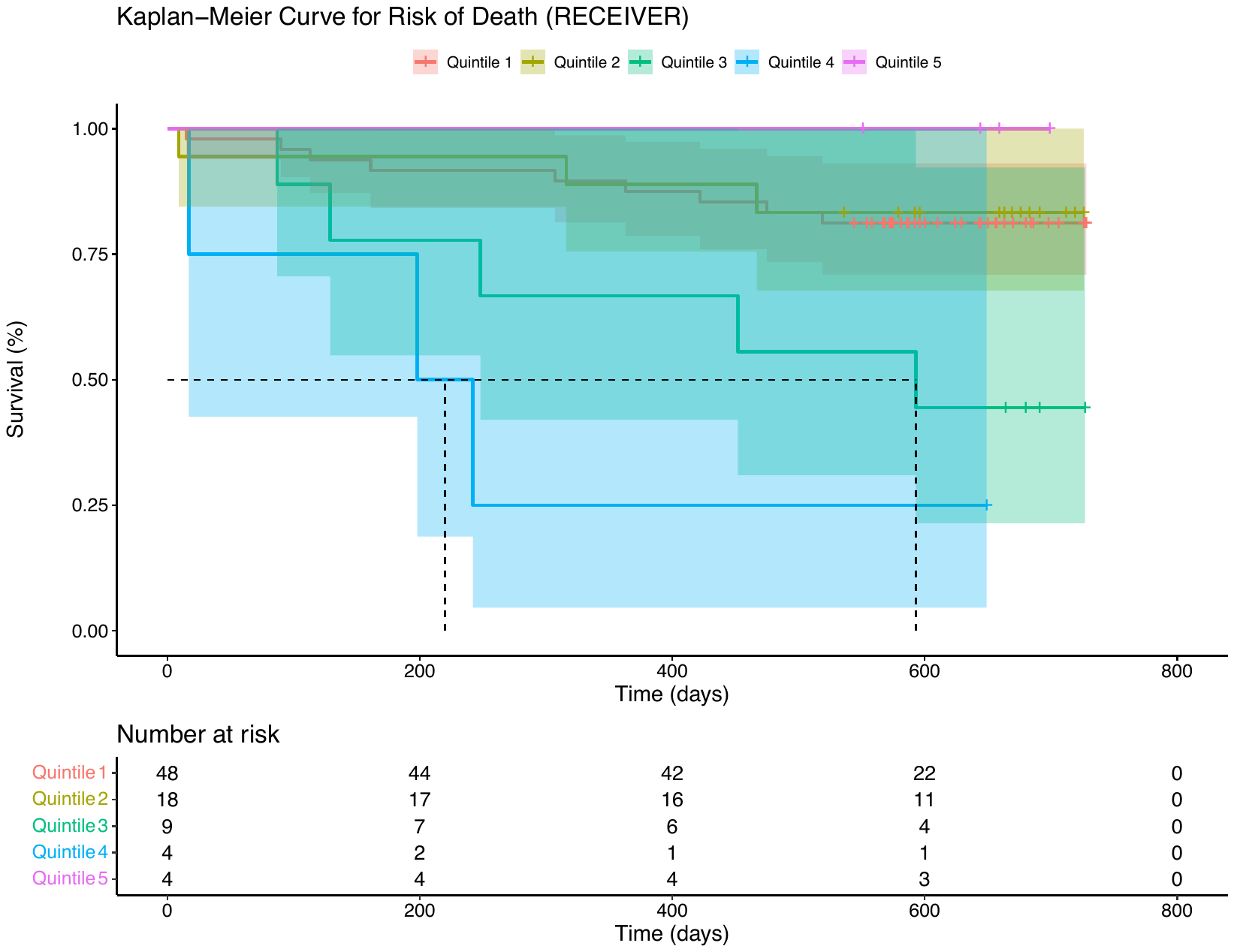

**Supplementary Figure 4** Kaplan-Meier survival plots showing no significant difference in time to admission or time to death from onboarding date until 31st August 2021 in RECEIVER trial participants stratified by SIMD quintile.

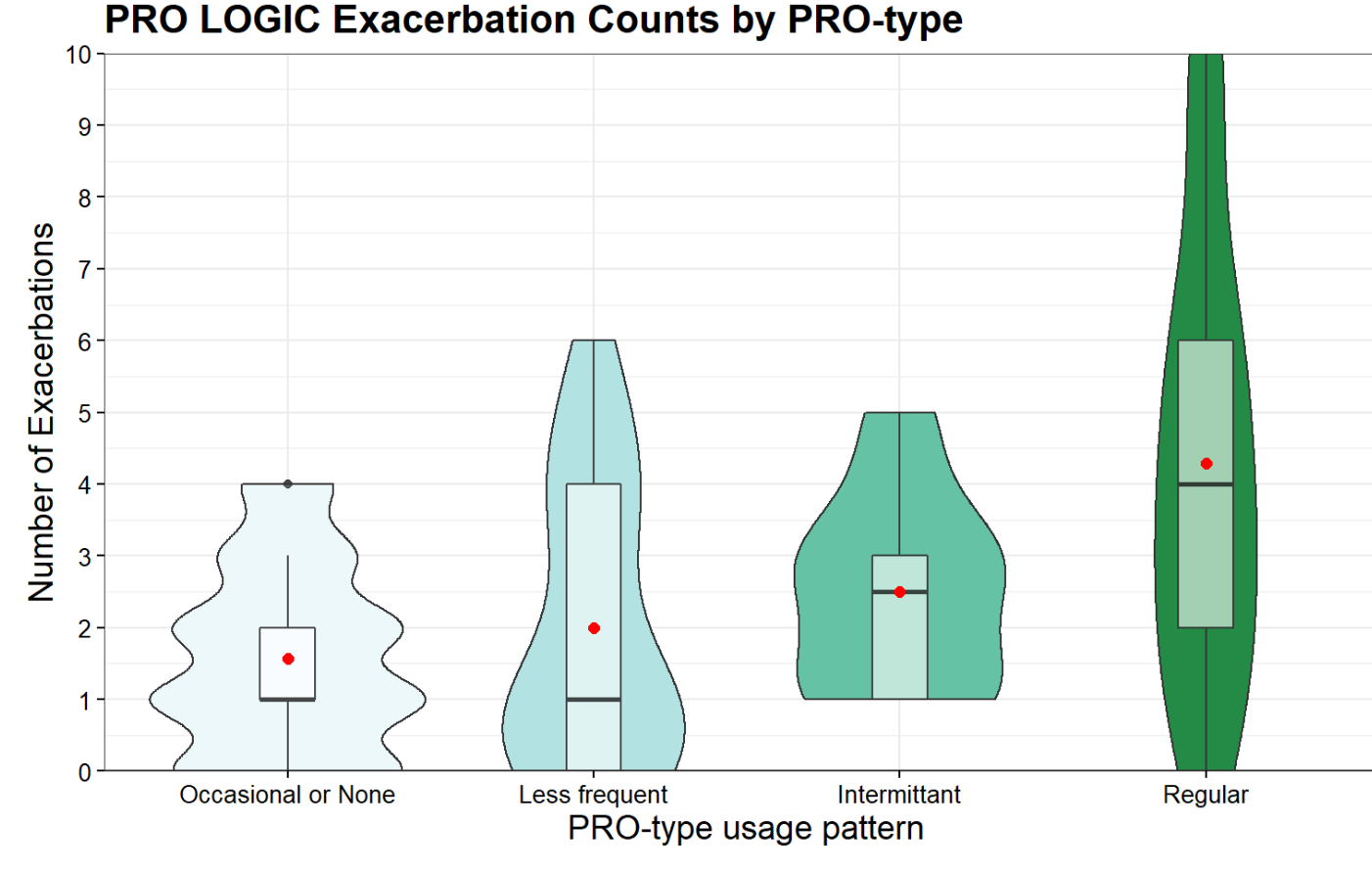
**Supplementary Figure 5** Violin boxplots summarising community-managed (clinician-assessed or self-treated) exacerbation events captured in the LenusCOPD patient app in the year following onboarding in the RECEIVER cohort subdivided subdivided by PRO-completion quartiles. Detection of a proportion of community-managed COPD exacerbations may be missed in participants who are not using the patient app regularly. Alternatively, patients with more frequent exacerbations may be more motivated to use the patient app regularly. See Figure 3C for PRO-completion data.

Red dot = median. Horizontal bar = mean.

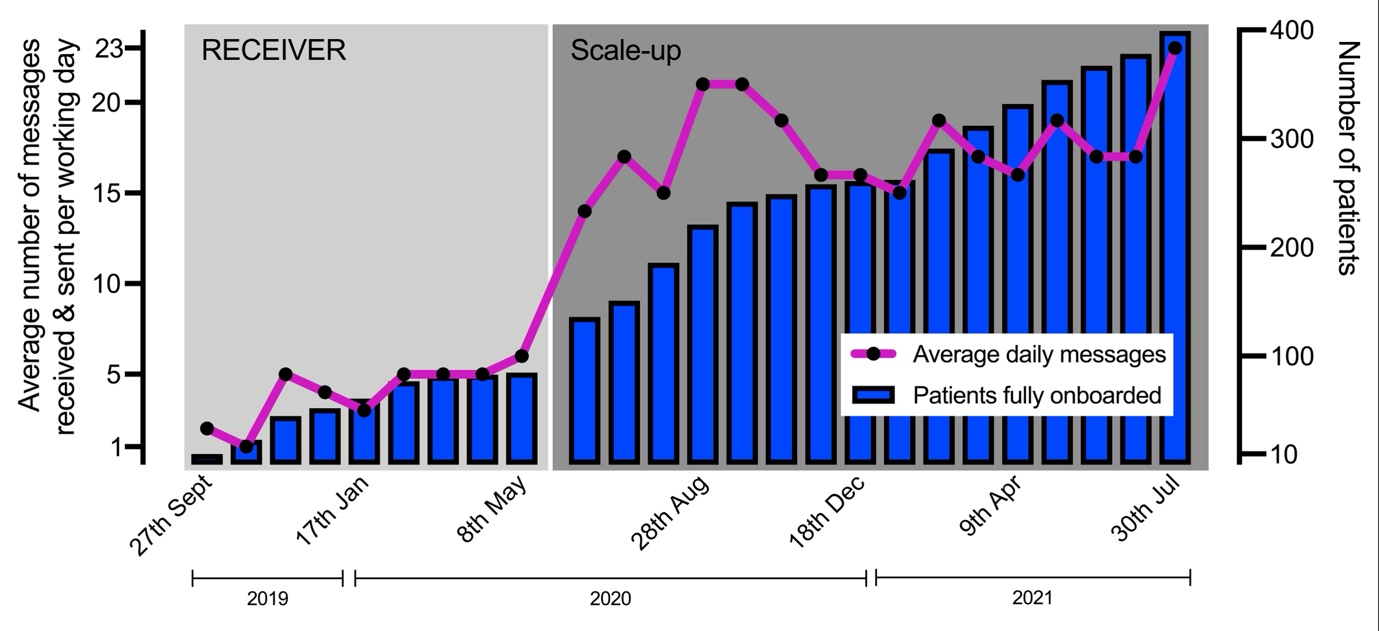

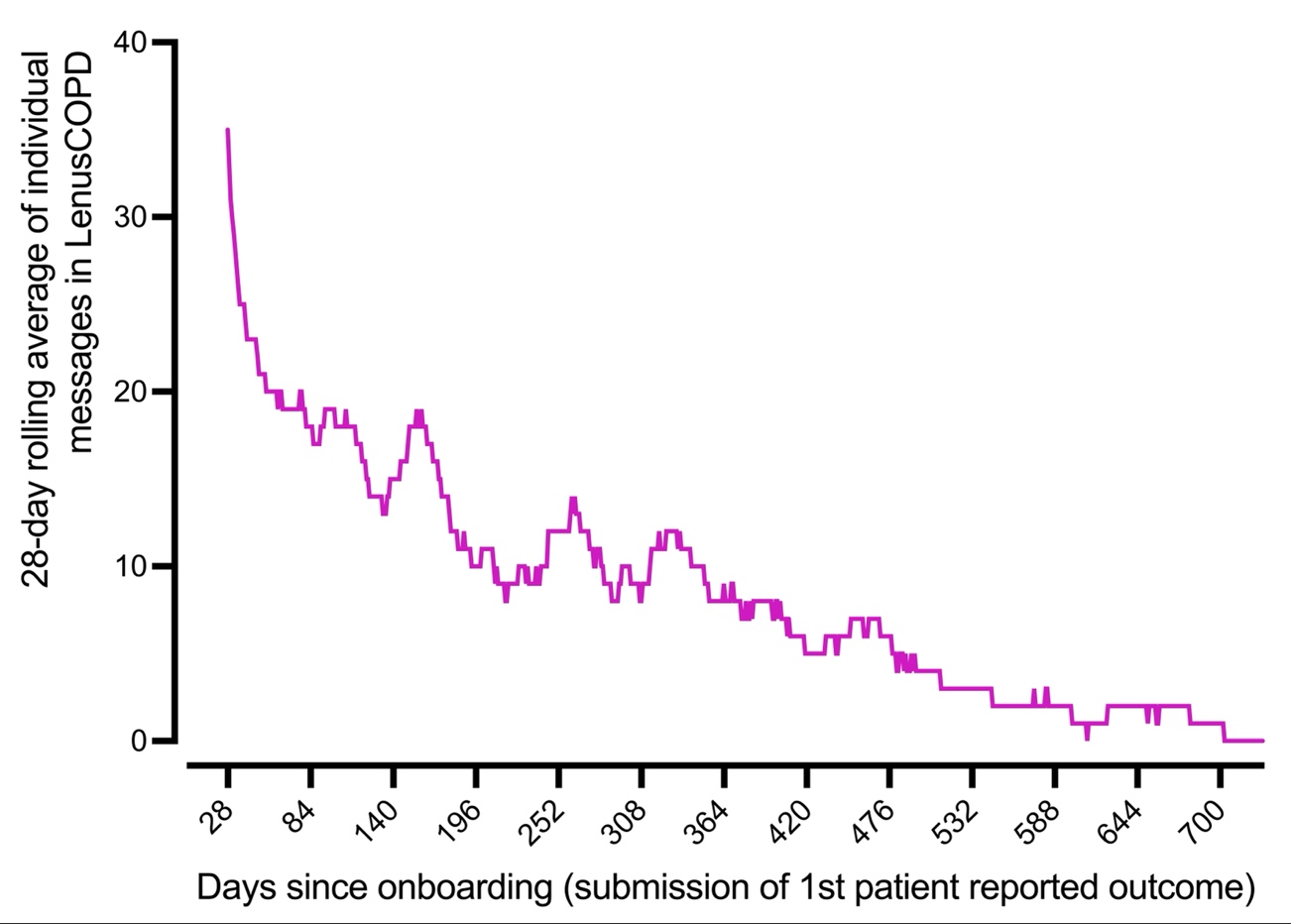

**Supplementary Figure 6** Patient messaging

Patient messaging volume initially increased with patient recruitment, then plateaud. Higher volume of patient<>clinician messages are sent in the 3 months following onboarding. Reduction in messaging activity is typical for self-management and telehealth interventions, potentially reflecting patient confidence with self-management and stabilisation of their condition. A higher number of messages and conversations were managed on Mondays, reflecting accumulation from weekend submissions. These metrics also informs the clinician resource required to adopt this service.
